# Relating in vitro neutralisation level and protection in the CVnCoV (CUREVAC) trial

**DOI:** 10.1101/2021.06.29.21259504

**Authors:** Deborah Cromer, Arnold Reynaldi, Megan Steain, James A Triccas, Miles P Davenport, David S Khoury

## Abstract

A recent study analysed the relationship between neutralising antibody response and protection from SARS-CoV-2 infection across eight vaccine platforms^1^. The efficacy results from a phase 2b/3 trial of a ninth vaccine candidate, CVnCoV (CUREVAC), was announced on 16 June 2021^2^. The low efficacy of this new mRNA vaccine, which showed only 47% protection from symptomatic SARS-CoV-2 infection, was surprising given the high efficacy of two previous mRNA-based vaccines^3,4^. A number of factors have been suggested to play a role in the low efficacy in the CVnCoV study, particularly around the dose and immunogenicity of the vaccine (which uses an unmodified mRNA construct^5,6^) and the potential role of infection with SARS-CoV-2 variants (which were the dominant strains observed in the CVnCoV trial)^2^.

## Main text

To investigate the potential effects of dose and immunogenicity in the CVnCoV construct we extracted data on in vitro neutralisation titre for three reported mRNA vaccines, CVnCoV^6^, mRNA-1273^7^, and BNT162b2^8^. To allow comparison of neutralisation levels between studies we normalised to the average convalescent titre in the same study (recognising that convalescent groups were not standardised between studies). Figure 1a compares dose and neutralisation levels across the 3 vaccines and suggests that the lower neutralisation in the CVnCoV study is consistent with the neutralisation observed when lower doses of mRNA-1273^7^, and BNT162b2^8^ were administered.

**Figure 1:**
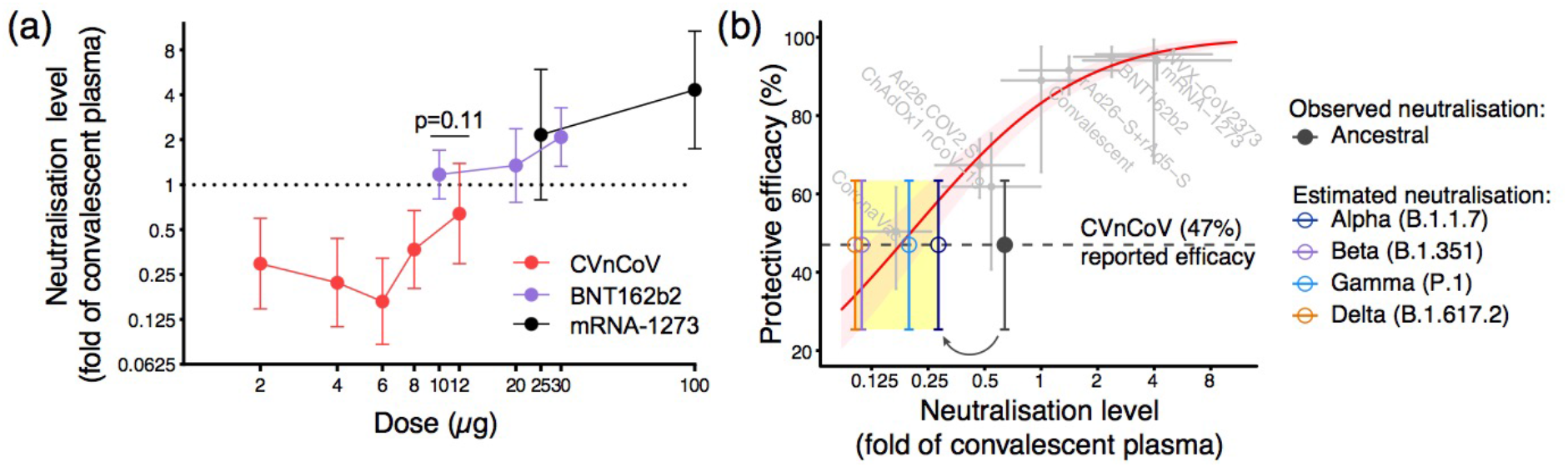
Potency, immunogenicity, and protection from SARS-CoV-2 infection. (a) The relationship between immunisation dose (x-axis) and in vitro neutralisation titre (expressed as a fold-change over convalescent serum in the same study)(y-axis) is shown for three mRNA-based vaccines, CVnCoV^6^, mRNA-1273^7^, and BNT162b2^8^. The neutralisation level of the 12 μg dose of CVnCoV was not significantly different from that of the 10 μg BNT162b2 dose (p=0.11, t-test). (b) The previously reported relationship between neutralisation level (normalised to the mean convalescent titre in the same study) and protection from symptomatic SARS-CoV-2 infection is shown in red^1^. The observed mean neutralisation level against ancestral SARS-CoV-2 virus in vitro (grey)^6^, as well as the predicted drop in neutralisation level (indicated by arrow and coloured dots and whiskers) against the alpha, beta, delta and gamma variants^10,11^ are plotted against the protective efficacy observed in the CVnCoV trial^2^.

Another factor suggested to affect the observed vaccine efficacy was the circulating SARS-CoV-2 variant viruses encountered during the CVnCoV trial. In the primary phase 3 studies used for licensure of the other eight vaccines, the ancestral virus (which matches the spike protein used as the vaccine immunogen) was the dominant strain in circulation^1^. However, more recent non-randomised studies have suggested a reduced efficacy of some of these vaccines against SARS-CoV2 variants^9^. In vitro studies have shown that many SARS-CoV-2 variants show a significant reduction in neutralisation titre compared to the ancestral virus, and that this effect is observed using serum from both convalescent and vaccinated subjects^10,11^. In the CVnCoV phase 3 trial the infecting virus was composed almost entirely of a variety of circulating SARS-CoV-2 variants. For example, the alpha (B.1.1.7) variant represented about 41% of infections in the CVnCoV trial and has been shown by others to have a 2.3-fold drop in neutralising titre compared to the ancestral virus for another mRNA vaccine^10^. Similarly, the beta (B.1.351), gamma (P.1) and delta (B.1.617.2) variants have been shown to have a 5.8, 3.2-fold and 6.3-fold drop in neutralising titre respectively^10,11^.

To visualise the potential effects of reduced neutralising level on vaccine efficacy, Figure 1b plots the in vitro neutralising titre (normalised to the mean convalescent sera) against the ancestral virus observed in the CVnCoV phase 1 / 2 study^6^ along with the observed efficacy in the Phase 3 trial. It also shows the predicted effects of the drop in neutralisation titre for the different variants listed above^10,11^. The observed efficacy against variants appears consistent with the initial level of neutralisation against the ancestral virus and the expected drop in neutralisation titre to variants (as reported for other mRNA vaccines).

This analysis suggests that both a lower dose than the other mRNA vaccines (Fig 1a), as well as the effects of SARS-CoV-2 variants in reducing the neutralising ability of vaccine-induced serological responses (Fig 1b) were significant contributors to the low efficacy observed in the CVnCoV study.

## Data Availability

All data and code are freely available from the corresponding author upon request.

## Ethics statement

This work was approved under the UNSW Sydney Human Research Ethics Committee (approval HC200242).

## Authorship Statement

All authors contributed to the data collection, design of the study, writing of the manuscript and revision of the manuscript. DSK, DC, AR, and MPD contributed to the modelling and statistical analysis of the data.

## Data Availability Statement

All data and code are freely available from the corresponding author upon request.

## Acknowledgments

The authors wish to thank Sarah Sasson for helpful comments and careful reading of the manuscript.

